# Early impact of school closure and social distancing for COVID-19 on the number of inpatients with childhood non-COVID-19 acute infections in Japan

**DOI:** 10.1101/2020.11.18.20233957

**Authors:** Kenji Kishimoto, Seiko Bun, Jung-ho Shin, Daisuke Takada, Tetsuji Morishita, Susumu Kunisawa, Yuichi Imanaka

**Author notes:** **Corresponding author:** Yuichi Imanaka, Department of Healthcare Economics and Quality Management, Graduate School of Medicine, Kyoto University, Yoshida Konoe-cho, Sakyo-ku, Kyoto 606-8501, Japan.

## Abstract

Many countries have implemented school closures as part of social distancing measures intended to control the spread of coronavirus disease 2019 (COVID-19). The aim of this study was to assess the early impact of nationwide school closure (March-May 2020) and social distancing for COVID-19 on the number of inpatients with major childhood infectious diseases in Japan. Using data from the Diagnosis Procedure Combination system in Japan, we identified patients aged 15 years or younger with admissions for a diagnosis of upper respiratory tract infection (URTI), lower respiratory tract infection (LRTI), influenza, gastrointestinal infection (GII), appendicitis, urinary tract infection (UTI), or skin and soft tissue infection (SSTI) between July 2018 and June 2020. Two periods were considered in the analysis: a pre- and a post-school-closure period. Changes in the trend of the weekly number of inpatients between the two periods were assessed using interrupted time-series analysis. A total of 75,053 patients in 210 hospitals were included. We found a marked reduction in the number of inpatients in the post-school-closure period, with an estimated reduction of 581 (standard error 42.9) inpatients per week (*p* < 0.001). The main part of the reduction was for pre-school children. Remarkable decreases in the number of inpatients with URI, LRTI, and GII were observed, while there were relatively mild changes in the influenza, appendicitis, UTI, and SSTI groups. We confirmed a marked reduction in the number of inpatients with childhood non-COVID-19 acute infections in the post-school-closure period.

## Introduction

The current coronavirus disease 2019 (COVID-19) pandemic [1,2] has led to over 900,000 deaths worldwide [3], and has resulted in substantial changes in the practice of health care. Most countries have implemented social distancing measures to limit the spread of the novel coronavirus SARS-CoV-2, which is responsible for COVID-19 [2,4]. Furthermore, many countries have implemented school closures nationwide as part of social distancing measures, although the effect of school closures on the epidemic has not been fully investigated [5-7]. In Japan, the first COVID-19 case was reported on January 16, 2020. As the severity of the outbreak became evident, the Japanese government called for the closure of all elementary schools (for children aged 6-12), junior high schools (for ages 12-15 years), and high schools (for ages 15-18 years) on February 27, 2020. Nursery schools were excluded from the nationwide closure request. A state of emergency was then declared, initially for seven prefectures on April 7, later nationwide on April 16, 2020. The state of emergency was lifted nationwide on May 25, 2020, and most schools had reopened by the first week of June 2020 [8].

The impact of the COVID-19 pandemic on health care for children has been described in areas such as emergency departments [9,10] and chronic disease care facilities [11,12]. The number of children with acute infection, the most common presentation of children to hospitals, also appears to be affected by the COVID-19 pandemic. One recent study showed a large decrease in pediatric emergency visits and admissions following emergency visits for viral infections after the school closure and national lockdown [13]. Another study has reported an unprecedented decline in hospital admissions for children younger than three years with respiratory infections after social distancing [14]. However, there is little information on the overall change in the number of hospitalized children with acute infections caused by school closure and social distancing. The aim of this study was to assess the early impact of school closure and social distancing for COVID-19 on the number of inpatients with major childhood infectious diseases in Japan using administrative claims data.

## Material and methods

### Data source

Data were obtained from the database of the Quality Indicator/Improvement Project (QIP), an ongoing project in Japan intended to monitor and improve clinical performance in acute care hospitals through the analysis of administrative claims data [15]. Over 500 hospitals across Japan, including both public and private hospitals, have participated in the QIP since its inception in 1995. The QIP database contains Diagnosis Procedure Combination (DPC) administrative claims data from the participating hospitals. The DPC data includes information on hospital codes, patient demographics, admission and discharge dates, admission routes, outcomes, primary and secondary diagnoses based on International Classification of Diseases 10th revision (ICD-10) codes, comorbidities, complications, and claims for medical services [16].

### Study population and disease definitions

Data from 257 hospitals in the QIP that had submitted DPC data on both inpatients and outpatients for the study period of July 2018 through June 2020 were used to select the study population. (Only data submitted prior to August 26, 2020, were considered.) Seven groups of infectious diseases were distinguished: upper respiratory tract infection (URTI), lower respiratory tract infection (LRTI), influenza, gastrointestinal infection (GII), appendicitis, urinary tract infection (UTI), and skin and soft tissue infection (SSTI). All patients aged 15 years or younger with admissions for a primary diagnosis and the most medical resource-intensive diagnosis within the seven disease groups between July 2018 and June 2020 were included in the study. Patients with a diagnosis of coronavirus infection or COVID-19 (defined by ICD-10 codes B34.2 or U07.1) were excluded. Two age groups were defined: pre-school children (aged 0-5 years) and school-age children (aged 6-15 years). URTI was defined by ICD-10 codes J00.x through J60.x. LRTI was defined by ICD-10 codes J12.x through J40.x. Influenza was defined by ICD-10 codes J09.x through J11.x. GII was defined by ICD-10 codes A00.x through A09.x. Appendicitis was defined by ICD-10 codes K35.x through K37.x. UTI was defined by ICD-10 codes N10.x, N13.6, N15.1, N30.x, N34.x, and N39.0. SSTI was defined by ICD-10 codes L00.x through L08.x.

### Statistical analysis

The primary outcome of interest was the weekly number of inpatients. We used the date of discharge rather than the date of admission for comparing the number of weekly inpatients, since the DPC data were generated after discharge. The secondary outcome was the change in the trend of the weekly number of inpatients before and after the school closure in March 2020. Two periods were considered in the analysis: the pre-school-closure period (July 1, 2018, through February 29, 2020) and the post-school-closure period (March 1, 2020, through June 30, 2020). Changes in the trend of the weekly number of inpatients were analyzed by segmented regressions and an interrupted time-series design to estimate the effects of the school closure [17]. The seasonality of hospital admissions for pediatric infectious disease was taken into account by including harmonic terms with a 52-week period. The intervention effect (decrease in the number of inpatients) was estimated by comparing estimates in the post-school-closure period to expected estimates from a Gaussian regression model. The validity of the regression model was assessed by visual inspection. Overall analysis and stratified analyses by age group and disease group were performed. A *p*-value of less than 0.05 was considered statistically significant; all tests were two-tailed. All analyses were conducted using R version 4.0.2 (The R Development Core Team, Vienna, Austria).

### Ethical considerations

This study was approved by the Ethics Committee, Graduate School of Medicine, Kyoto University (approval number: R0135), and conducted in accordance with the Ethical Guidelines for Medical and Health Research Involving Human Subjects of the Ministry of Health, Labour and Welfare, Japan.

## Results

In all, 75,101 patients in 210 hospitals who met the inclusion criteria during the study period were identified from the database. Forty-eight patients with coronavirus infection or COVID-19 were excluded from the group. Thus, 75,053 patients in 210 hospitals in the seven disease groups were included in the analyses. Patient characteristics are summarized in Table 1. LRTI was the most common disease group (51.9%). The second most frequent group was GII (15.4%), followed by URTI (13.8%), influenza (5.8%), UTI (5.1%), appendicitis (4.7%) and SSTI (3.3%).

**Table 1.**
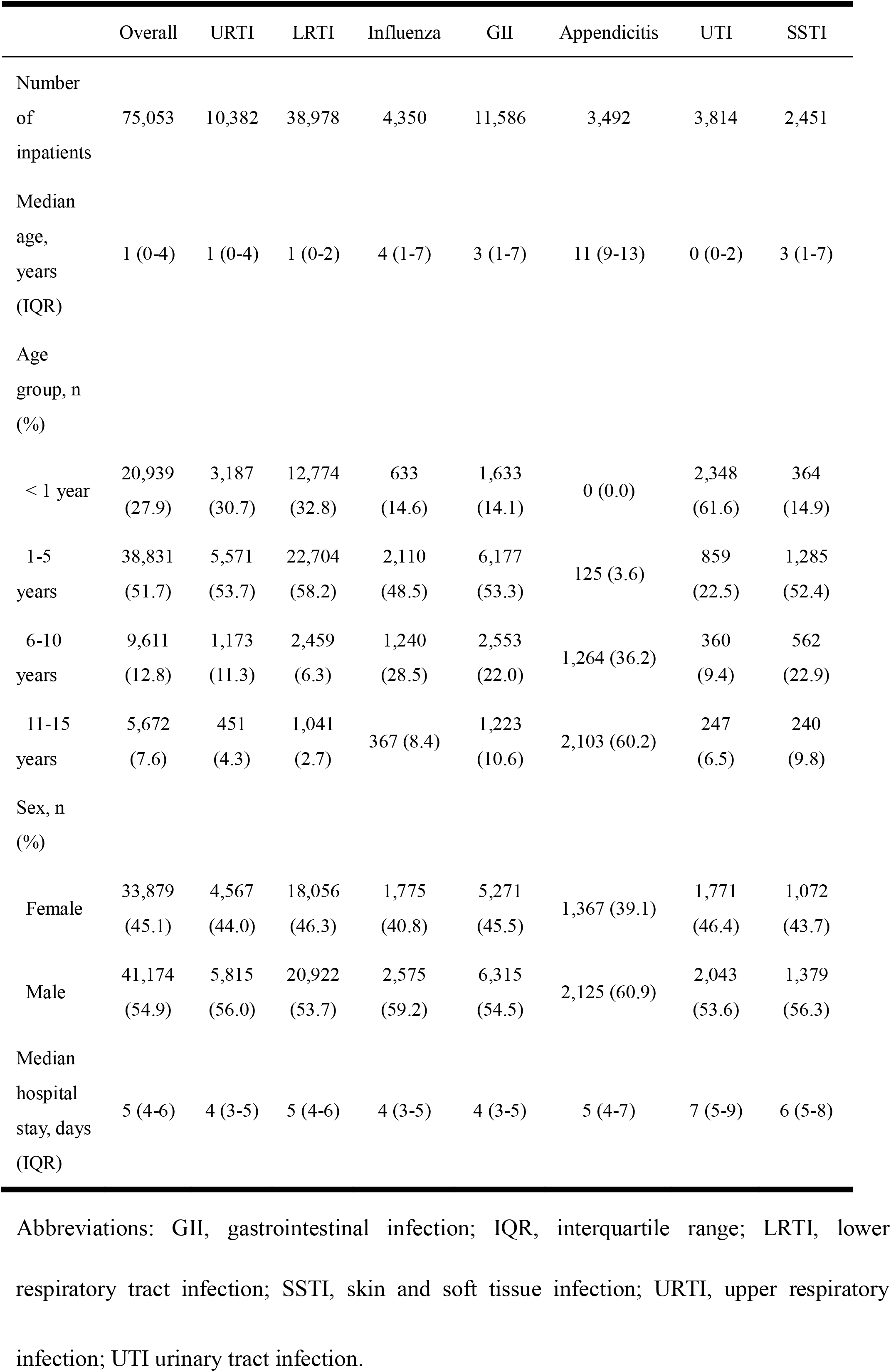
Characteristics of patients by overall inpatients and disease groups

|  | Overall | URTI | LRTI | Influenza | GII | Appendicitis | UTI | SSTI |
| --- | --- | --- | --- | --- | --- | --- | --- | --- |
| Number of inpatients | 75,053 | 10,382 | 38,978 | 4,350 | 11,586 | 3,492 | 3,814 | 2,451 |
| Median age, years (IQR) | 1 (0-4) | 1 (0-4) | 1 (0-2) | 4 (1-7) | 3 (1-7) | 11 (9-13) | 0 (0-2) | 3 (1-7) |
| Age group, n (%) |  |  |  |  |  |  |  |  |
| < 1 year | 20,939 (27.9) | 3,187 (30.7) | 12,774 (32.8) | 633 (14.6) | 1,633 (14.1) | 0 (0.0) | 2,348 (61.6) | 364 (14.9) |
| 1-5 years | 38,831 (51.7) | 5,571 (53.7) | 22,704 (58.2) | 2,110 (48.5) | 6,177 (53.3) | 125 (3.6) | 859 (22.5) | 1,285 (52.4) |
| 6-10 years | 9,611 (12.8) | 1,173 (11.3) | 2,459 (6.3) | 1,240 (28.5) | 2,553 (22.0) | 1,264 (36.2) | 360 (9.4) | 562 (22.9) |
| 11-15 years | 5,672 (7.6) | 451 (4.3) | 1,041 (2.7) | 367 (8.4) | 1,223 (10.6) | 2,103 (60.2) | 247 (6.5) | 240 (9.8) |
| Sex, n (%) |  |  |  |  |  |  |  |  |
| Female | 33,879 (45.1) | 4,567 (44.0) | 18,056 (46.3) | 1,775 (40.8) | 5,271 (45.5) | 1,367 (39.1) | 1,771 (46.4) | 1,072 (43.7) |
| Male | 41,174 (54.9) | 5,815 (56.0) | 20,922 (53.7) | 2,575 (59.2) | 6,315 (54.5) | 2,125 (60.9) | 2,043 (53.6) | 1,379 (56.3) |
| Median hospital stay, days (IQR) | 5 (4-6) | 4 (3-5) | 5 (4-6) | 4 (3-5) | 4 (3-5) | 5 (4-7) | 7 (5-9) | 6 (5-8) |

Abbreviations: GII, gastrointestinal infection; IQR, interquartile range; LRTI, lower
respiratory tract infection; SSTI, skin and soft tissue infection; URTI, upper respiratory infection; UTI urinary tract infection.

### Change in the overall number of inpatients

Figure 1 illustrates the time trend of weekly number of inpatients and the results of the interrupted time-series analysis. We found a marked reduction in the number of inpatients in the post-school-closure period (Figure 1a). The reduction in the number of inpatients had already become substantial by the end of March 2020. The overall weekly number of inpatients was 638 in the last week of February 2020, 375 (−41.2%) in the last week of March 2020, 276 (−56.7%) in the last week of April 2020, and 166 (−74.0%) in the last week of May 2020. The estimated impact from the interrupted time series analysis was a reduction of 581 (standard error 42.9) inpatients per week between the two periods (*p* < 0.001).

**Figure 1.**
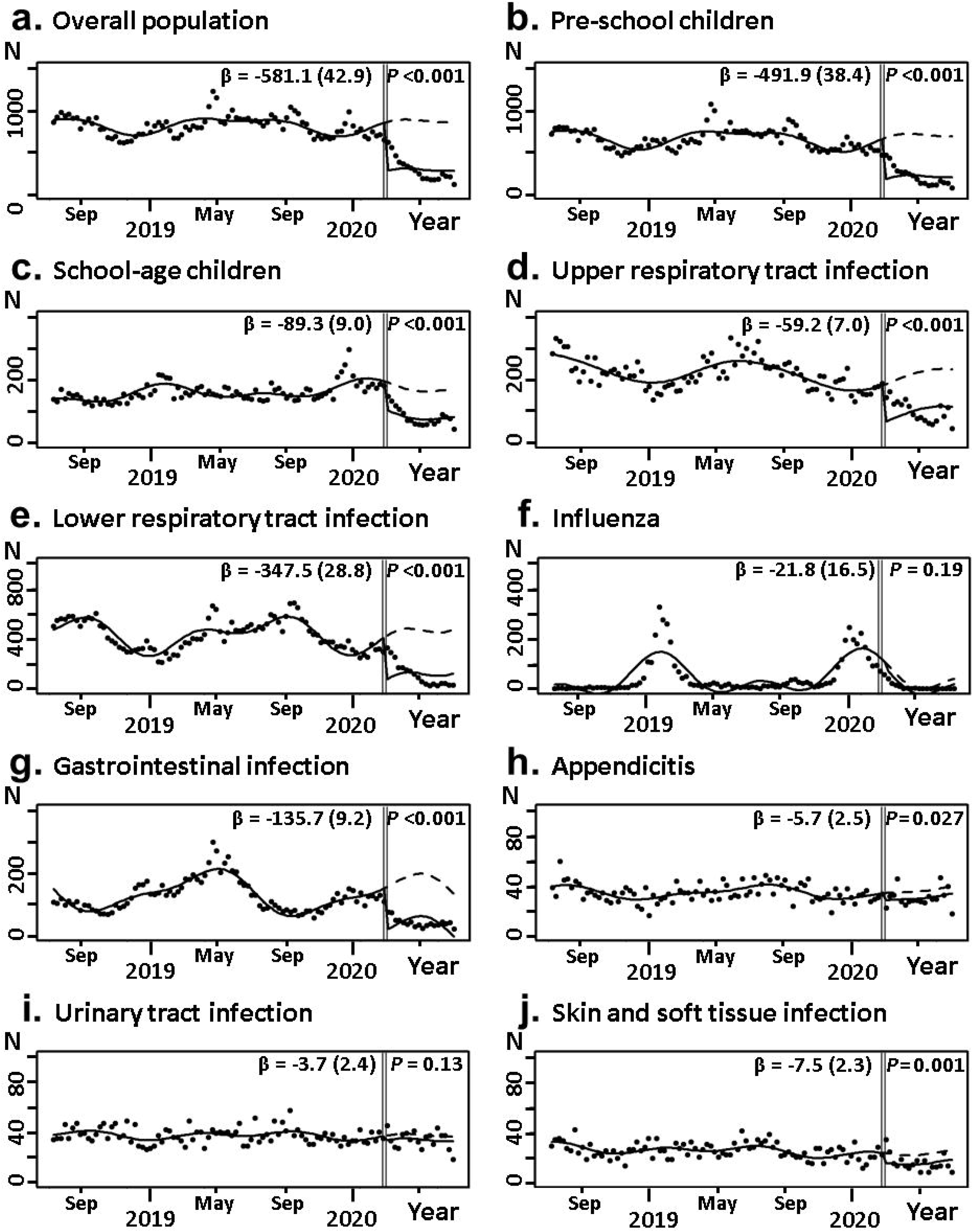
Time trend in the number of inpatients between July 2018 and June 2020; solid lines indicate observed trend following school closure for COVID-19, and dashed lines indicate predicted trend. The estimated coefficient for the change in the number of inpatients per week (standard error) and *p*-value are shown in each analysis. Vertical double bars indicate the initiation of nationwide school closure in March 2020. (a) Overall population; (b) pre-school children (aged 0-5 years); (c) school-age children (aged 6-15 years); (d) patients with URTI; (e) patients with LRTI; (f) patients with influenza; (g) patients with GII; (h) patients with appendicitis; (i) patients with UTI; (j) patients with SSTI. Abbreviations: COVID-19, coronavirus disease 2019; GII, gastrointestinal infection; LRTI, lower respiratory tract infection; SSTI, skin and soft tissue infection; URTI, upper respiratory infection; UTI urinary tract infection.

### Stratified analysis by age group and disease group

A major reduction in the number of inpatients was observed in the post-school-closure period for both pre-school children and school-age children (Figure 1b and 1c). The weekly number of pre-school inpatients in the last week of March 2020 was 41.3% lower than in the last week of February 2020. The result for school-age children was similar; for this group, the weekly number of inpatients in the last week of March 2020 was 41.1% lower than in the last week of February 2020. As the baseline numbers of inpatients was higher for pre-school children than for school-age children, pre-school children accounted for the main part of the reduction in the number of inpatients. The interrupted time series analysis confirmed a prominent decrease in the number of inpatients in both age groups. We found that the magnitude of the decrease in the number of inpatients after the school closure was different between the disease groups (Figure 1d-1j). Remarkable decreases in the number of inpatients with URI, LRTI, and GII were observed in the post-school-closure period. By contrast, there were relatively mild changes in the influenza, appendicitis, UTI, and SSTI groups. The main part of the reduction in the number of inpatients was in the LRTI and GII groups. The interrupted time series analysis showed a pronounced decrease in the number of inpatients in the URTI, LRTI, and GII groups.

## Discussion

We found that the number of children hospitalized as inpatients for non-COVID-19 acute infections decreased markedly in the post-school-closure period in Japan. This was true for both pre-school children and school-age children. We also found the different impact on the number of inpatients between the disease groups. These results suggest the potential impact of school closure and social distancing for COVID-19 on the number of inpatients with childhood infectious diseases. We considered the nationwide school closure rather than the government’s declaration of a state of emergency as the primary cause of the reduction, as the reduction had already been observed in March 2020, prior to the emergency declaration in April 2020.

Our results showed that the decrease in the number of inpatients after the school closure was observed not only in school-age children but also in pre-school children, which is consistent with previous reports on the effect of the COVID-19 pandemic on pediatric acute infection [13,14]. Indeed, the absolute decrease in the number of inpatients was much greater in pre-school children. Moreover, the reduction in the number of inpatients was more prominent in transmissible infectious diseases such as URI, LRTI, and GII. Regarding admissions for influenza, the relatively mild change in the post-school-closure period may be due to its coinciding with the end of the influenza season. While these findings could be used to support a hypothesis that the decreased transmission of pathogens among younger children due to social distancing following school closure is responsible for the remarkable decrease in the number of inpatients observed in this study, another possible explanation is hospital avoidance during the COVID-19 pandemic [18,19]. This would not be without precedent as previous studies have reported healthcare avoidance in epidemics or outbreaks of emerging infectious diseases [20,21]. Further studies using community-based data that include primary care settings are needed to estimate the impact of hospital avoidance. As UTI has been regarded as a non-transmissible disease in past studies of childhood infection [22,23], the fact that the number of inpatients for UTI did not show an obvious change in the post-school-closure period may suggest that the impact of hospital avoidance was not particularly high.

Current evidence on the effect of school closures and social distancing has been derived almost entirely from studies of influenza outbreaks [24,25]. The effectiveness of social distancing among children in other outbreaks, including COVID-19, remains unclear. Dealing with the COVID-19 pandemic clearly requires massive healthcare resources [26,27]. A reduction in hospital admissions for childhood infections may save healthcare resources that could then be utilized for COVID-19 care. As the estimated costs and negative impacts of school closure are significant [28-30], our results can be useful for policy making in the ongoing COVID-19 pandemic or in future outbreaks.

This study has several limitations. The study population, which was restricted to a certain segment of hospitals in the QIP, is perhaps the primary limitation. The movement of patients from the enrolled hospitals to other facilities during the COVID-19 pandemic may have caused an overestimation of the reduction in the number of inpatients in the post-school-closure period. However, a relatively consistent trend in the number of inpatients in the influenza and UTI groups during the study period suggests that there was no large-scale movement of patients from the enrolled hospitals. A lack of information on disease severity is another limitation of the study. Changes in disease severity in the enrolled patients between the two periods were not assessed, as the DPC data are not sufficient to evaluate the severity of a childhood infectious disease. A third limitation is that the study included only the Japanese population. It should be noted that no mandatory social distancing or so-called lockdowns were imposed by the Japanese government. As there have been considerable regional differences in the spread of COVID-19 and in the various epidemic-control measures taken globally, it is difficult to predict whether the present findings for Japan are applicable elsewhere. Despite these limitations, our study provides novel information on the impact of the COVID-19 pandemic on health care for children.

In summary, we confirmed a marked reduction in the number of school-age and pre-school inpatients for non-COVID-19 acute infections in the post-school-closure period. We showed that the main part of the reduction was for pre-school children, particularly in the LRTI and GII disease groups.

## Data Availability

The datasets analyzed during the current study available from the corresponding author on reasonable request.

## Funding

This study was supported by the Japan Society for the Promotion of Science [JSPS KAKENHI Grant Number JP19H01075 to Y.I.] and GAP Fund Program of Kyoto University. Acknowledgement

We gratefully acknowledge the participating hospitals in the QIP and their staffs. We also acknowledge the colleagues in the Department of Healthcare Economics and Quality Management, Graduate School of Medicine, Kyoto University, Japan.

